# Analgesic Efficacy of Native Himalayan Shilajit as Add-On Therapy in Myofascial Pain Syndrome: An Exploratory Pilot Clinical Trial

**DOI:** 10.64898/2026.07.24.26357716

**Authors:** Darshan Basavaraja, Ravi Kant, Venkatesh S Pai, Rajkumar Yadav, Gaurav Chikara, Shailly Tomar, Debabrata Sircar, Katkar Rahul Sambhaji, Prasan Kumar Panda

## Abstract

**BACKGROUND:** Myofascial Pain Syndrome (MPS) is a common musculoskeletal pain condition associated with myofascial trigger points that has been reported to occur in 30-93% of patients who present with musculoskeletal pain. The current pharmacologic treatments (such as nonsteroidal anti-inflammatory drugs [NSAIDs], muscle relaxants, and tricyclic antidepressants) are only partially effective and have side effects. Shilajit, a mineral-organic exudate from the Himalayas, has antioxidant, anti-inflammatory, mitochondrial bioenergetic, and central analgesic effects and has not previously been examined as an analgesic in any musculoskeletal pain condition.

**METHODS:** This was an exploratory pilot clinical trial with open-label design in a single arm for an 18-month period at All India Institute of Medical Sciences (AIIMS), Rishikesh, India. Patients aged 18 to 65 years with clinically diagnosed MPS (Simons et al. 1999 criteria) and a baseline visual analog scale (VAS) score >4 were enrolled. Native Himalayan Shilajit 250 mg daily was administered as add-on therapy for 49 days. The main outcome was the percentage of participants with ≥30% VAS reduction at Day 49. The intensity of pain, the dose of analgesics consumed and the number of trigger points were evaluated at five time points (Day 0, 12, 24, 36, 49). Throughout, adverse events were monitored.

**RESULTS:** Of 80 enrolled participants, 76 (95.0%) completed the per-protocol analysis. Mean age was 41.25 ± 9.05 years; 52.6% were male. A total of 56 of 76 participants (73.7%; 95% CI: 62.1– 82.8%) achieved the primary endpoint. Mean VAS score declined from 6.63 ± 1.08 at baseline to.63 ± 1.72 at Day 49 (mean reduction 45.3%; Friedman χ^2^ = 278.5, p<0.001). The first signs of pain reduction were seen at Day 24. The number of analgesic doses consumed decreased by 75.5% during the study period (χ^2^ = 126.1, p<0.001). There was a significant reduction in trigger point count from baseline to Day 49 (p=0.031) of 23.4%. One Grade 2 adverse event (gastrointestinal irritation, Day 28, resolved within 24 hours, no drug discontinuation) occurred; no serious adverse events were reported. Trial registration: CTRI/2025/06/088636. The study was not funded by any external sources.

**CONCLUSIONS:** Native Himalayan Shilajit 250 mg/day for 49 days was associated with clinically and statistically significant reductions in pain intensity, analgesic consumption, and trigger-point burden in patients with MPS, with a favorable safety profile. These findings warrant confirmation in a larger, randomized, placebo-controlled trial.

## Background

Musculoskeletal disorders collectively contribute over 115 million disability-adjusted life years (DALYs) globally, with low back pain ranking among the top contributors to the worldwide burden of disease(1). Myofascial Pain Syndrome (MPS) is a highly prevalent localized condition which is often underdiagnosed and characterized by hyperirritable myofascial trigger points (MTrPs) in taut bands of skeletal muscle that are exquisitely tender on palpation and are able to produce characteristic referred pain patterns in predictable distributions(2–5).MPS is estimated to be present in about 30% of primary care pain consultations, and MTrPs are found in 30–93% of patients presenting with musculoskeletal pain(6–8).

The pathophysiology is based on the integrated hypothesis, in which persistent acetylcholine leakage at dysfunctional neuromuscular junctions results in a persistently shortened sarcomere, localized ischemia, and ultimately an energy crisis due to ATP depletion(4,9), leading to peripheral sensitization of muscle nociceptors, and with sustained nociceptive input, central sensitization at the dorsal horn, which makes the condition difficult to treat and with high recurrence rates and no universally effective therapy(9–11).

The current pharmacological treatment, such as nonsteroidal anti-inflammatory drugs (NSAIDs), muscle relaxants, tricyclic antidepressants, and trigger point injections, is only partially effective and often only temporarily effective, and has significant side effects including gastrointestinal, renal, cardiovascular, and dependence(12–15).

Shilajit, known in classical Ayurveda as Shilajatu (WHO ITA Term ID: ITA-7.2.2.6)(16) and in Western literature as mineral pitch or *Asphaltum punjabianum*, is a complex organo-mineral exudate that seeps from rock fissures in the Himalayan belt at altitudes of 1000–5000 meters(17– 19).

The main bioactive components of Shilajit—fulvic acid, dibenzo-alpha-pyrones (DBPs), humic acids, and ionic trace minerals(17,20)—converge on the core pathophysiological mechanisms of MPS: fulvic acid and DBPs act as mitochondrial electron carriers enhancing ATP synthesis and restoring adenylate energy charge, directly targeting the trigger point energy crisis(21,22); fulvic acid inhibits complement-mediated pro-inflammatory pathways and modulates TNF-α and interleukin signalling(23,24); Shilajit exhibits glycine- and GABA-mimetic activity at substantia gelatinosa neurons, thus providing central analgesic and sedative effects(25). Shilajit shows potent free radical scavenging activity, reducing the oxidative stress that perpetuates ischemia-reperfusion injury at trigger points(26). The analgesic effects have been demonstrated in animal models of thermal hyperalgesia(27), while several clinical trials in humans have shown a good safety profile at therapeutic doses(28–31). However, no clinical trial has yet assessed native Himalayan Shilajit as an analgesic in any musculoskeletal pain condition. We performed this exploratory pilot clinical trial to assess the analgesic efficacy, analgesic-sparing effect and safety of native Himalayan Shilajit in MPS patients at AIIMS, Rishikesh, India.

## Methods

### Study Design and Setting

This was an exploratory open-label, single-arm clinical trial over 18 months at the Department of General Medicine, in association with the Department of Physical Medicine and Rehabilitation (PMR), AIIMS Rishikesh, Uttarakhand, India. Subjects were selected from the outpatient clinics of both specialties. The study was approved by the Institutional Ethics Committee of AIIMS Rishikesh (Clearance No. AIIMS/IEC/25/273 dated 02/05/2025) and registered in the Clinical Trials Registry – India (CTRI/2025/06/088636). Written informed consent was obtained from all participants.

### Participants

Eligible adults were those aged 18 to 65 years who had clinically diagnosed MPS and a baseline visual analog scale (VAS) score >4. MPS diagnosis was confirmed based on the criteria of Simons et al. (1999)(32,33), which included all five major criteria (localized spontaneous pain, referred pain pattern, palpable taut band, hypersensitive MTrP, and restricted range of motion), and at least one minor criterion (reproduction of pain on MTrP compression, local twitch response, or relief with stretching). Pregnancy or lactation, chronic kidney disease or decompensated liver disease, presence of malignancy, use of anticoagulants, known hypersensitivity to Shilajit, severe psychiatric disorders, neurological disorders that affect pain perception, systemic inflammatory or rheumatological disease, and other clinical trials within the last 6 months were considered as exclusion criteria.

### Study Drug

Native Himalayan Shilajit was purified by classical Ayurvedic Śodhana (purification) process at Sivananda Ashram, Ganeshpur, Uttarkashi, Uttarakhand using the method of Triphala Kwath Śodhana (purification process) as described in the Rasaratna Samuccaya, which involves dissolving in water, filtering through various steps, and then cooking for a prolonged period with milk and Yogavahi bioenhancer (gomutra)(34,35). The purified material was analytically checked at the Department of Biosciences and Bioengineering, IIT Roorkee, to ensure that there was no presence of heavy metals and microbial contaminants. Tablets of 500 mg each with 250 mg purified Shilajit and 250 mg Triphala (as pharmaceutical binding agent) were prepared by Nagarjun Pharmacy, Dehradun. The dose of 250 mg was similar to that used in published human clinical trials(29,31), and used as add-on therapy, one tablet daily in the postprandial period for 49 days.

### Outcomes

The primary outcome was the proportion of participants who had a ≥30% reduction in VAS score from baseline to Day 49, which was based on the minimum clinically important difference for chronic pain trials as defined by Farrar et al. and recommended by IMMPACT(36,37). Pain was measured using a validated VAS (0–10 cm) with established reliability and validity in chronic musculoskeletal pain(38,39).

This was assessed at Day 0, Day 12, Day 24, Day 36, and Day 49. Secondary outcomes included safety (incidence, severity [Division of AIDS (DAIDS) scale] and causality [World Health Organization–Uppsala Monitoring Centre (WHO-UMC) scale]) of adverse events during the study period. Exploratory outcomes were: change in VAS score at each timepoint; change in active MTrP count; and the number of doses of analgesics taken per interval (Day 0–12, Day 12– 24, Day 24–36, Day 36–49). Patient self-report and pill count were used to evaluate treatment adherence (>95% for per-protocol analysis).

### Sample Size

The sample size was calculated using a prevalence-based finite population correction formula with the estimated annual MPS OPD caseload at AIIMS Rishikesh (N=250) as the reference population (conservative proportion p=0.50, margin of error d=0.10, 95% CI, Z=1.96) with an anticipated loss to follow-up of 10% added, resulting in n=70, which was then rounded up to n=80. This was a considerable number exceeding the published thresholds of pilot studies(40– 42).

### Statistical Analysis

The data were analyzed using IBM SPSS version 30.0 and entered in Microsoft Excel 2024. The Shapiro-Wilk test was performed to determine normality. Continuous variables are presented as mean ± SD or median (IQR) and categorical variables are presented as frequency and percentage (95% CI). Friedman test with Nemenyi post-hoc comparisons was used to determine the changes in VAS score between timepoints. The time to ≥30% VAS reduction was determined using Kaplan-Meier survival analysis, and subjects who did not achieve the criterion by Day 49 were censored. The overall trajectory was compared between the pain severity subgroups by Generalized Estimating Equations (GEE). Baseline predictors of treatment response (responder status: ≥30% VAS reduction) were evaluated by the Wilcoxon-Mann-Whitney U test, independent-samples t-test, Chi-squared test or Fisher’s exact test. Multivariable logistic regression was used to identify independent predictors. The Friedman test with Nemenyi comparisons was used for each of the following: Analgesic dose counts and trigger point changes. A p-value of <0.05 was deemed as statistically significant.

## Results

### Participant Flow and Baseline Characteristics

A total of 80 participants participated in the study, and 76 (95.0%) completed the per-protocol analysis through Day 49. Four participants (5.0%) were lost to follow-up after the baseline assessment despite repeated contact attempts. All four had attended the Day 0 visit but did not return for the Day 49 assessment.

The mean age was 41.25 ± 9.05 years (range 26–64); 52.6% were male. Most participants (81.6%) had no comorbidities; hypertension was the most common (13.2%). Mean BMI was 23.42 ± 2.50 kg/m^2^. The trapezius was the most frequently involved muscle (46.1%), followed by levator scapulae (21.1%) and rhomboids (19.7%). Mean pain duration was 3.91 ± 2.37 months. All participants had previously received analgesics. The mean baseline VAS score was 6.63 ± 1.08 (median 7.00, IQR 6.00–7.25), corresponding to 57 participants (75.0%) with moderate pain (VAS 3.5–7.4) and 19 (25.0%) with severe pain (VAS ≥7.5) per the classification of Boonstra et al(43). Baseline characteristics are summarised in Table 1.

**Table 1.**
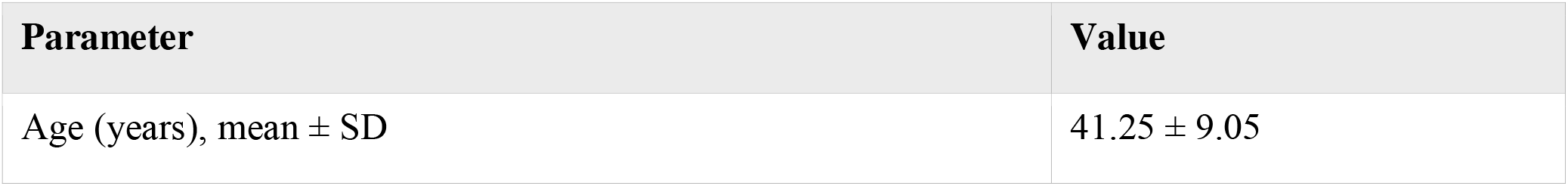
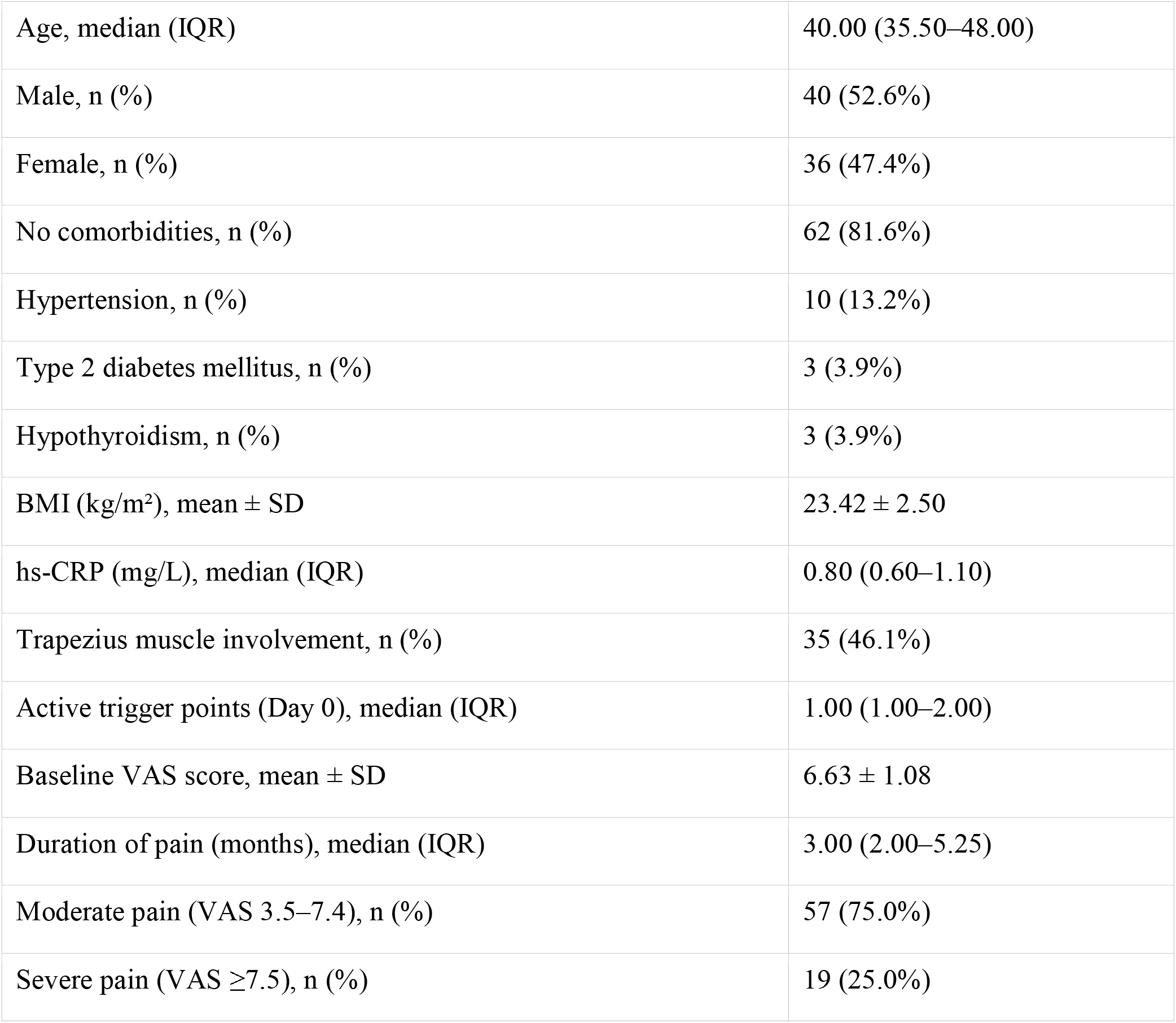
Baseline Demographics and Clinical Characteristics (Per-Protocol Analysis, n=76).

### primary Outcome: Clinically Significant Pain Reduction

Of 76 per-protocol participants, 56 (73.7%; 95% CI: 62.1–82.8%) achieved the primary endpoint of ≥30% VAS reduction by Day 49; 20 (26.3%; 95% CI: 17.2–37.9%) did not. Mean VAS declined progressively from 6.63 ± 1.08 at Day 0 to 3.63 ± 1.72 at Day 49, a mean percentage reduction of 45.3% (Friedman χ^2^ = 278.5, p<0.001). The difference between Day 0 and Day 12 was not statistically significant (mean difference 0.53 ± 0.58, p=0.168), thus demonstrating a delayed onset. The significant reduction first appeared at Day 24 (mean difference 1.62 ± 0.80, median percent reduction 33.3%, p<0.001), then there was a transient plateau from Day 24 to Day 36 (p=0.686), and a further significant reduction from Day 36 to Day 49 (mean difference 1.11 ± 0.62, p<0.001). Data for VAS at each time point are shown in Table 2.

**Table 2.** Change in VAS Score Across Study Timepoints (n=76).

| Timepoint | Mean (SD) | Median (IQR) | Range | P value vs. Day 0 |
| --- | --- | --- | --- | --- |
| Day 0 | 6.63 (1.08) | 7.00 (6.00–7.25) | 5.00–9.00 | — |
| Day 12 | 6.11 (1.15) | 6.00 (5.00–7.00) | 4.00–8.00 | 0.168 (NS) |
| Day 24 | 5.01 (1.28) | 5.00 (4.00–6.00) | 2.00–8.00 | <0.001 |
| Day 36 | 4.74 (1.41) | 5.00 (4.00–6.00) | 1.00–8.00 | <0.001 |
| Day 49 | 3.63 (1.72) | 4.00 (3.00–5.00) | 0.00–7.00 | <0.001 |

*Post-hoc pairwise comparisons by Nemenyi test. NS = not significant.*

### Kaplan-Meier Analysis: Time to Response

Clinically significant analgesic response (VAS reduction ≥30%) was estimated using the Kaplan-Meier survival analysis, and subjects who did not meet the criterion by the end of Day 49 were considered censored. The curve did not change from Day 0 to Day 24, indicating that none of the participants reached the response threshold within the first 24 days. The cumulative response rates were 30.3% (23 new responders) at Day 24, 39.5% (7 additional responders) at Day 36, and 73.7% (26 additional responders) at Day 49. The median time to response was Day 49 and the largest number of new responders was at the final assessment.

### Predictors of Treatment Response

Baseline VAS score was the only statistically significant independent predictor of response (OR 0.13, 95% CI: 0.02–0.56, p=0.02) as determined by multivariable logistic regression, with higher baseline pain severity being associated with lower likelihood of response. Older age was modestly associated with greater likelihood of response in univariable analysis (W=729.000, p=0.047; point-biserial r=0.19), but was not significant in the multivariable model. There were no significant associations with gender, BMI, hs-CRP, duration of pain, comorbidities, muscle group or baseline trigger point count. Both moderate (VAS 3.5-7.4) and severe (VAS ≥7.5) subgroups showed statistically significant within-group reductions (Friedman χ^2^=210.6 and χ^2^=68.2 respectively, p<0.001 for both), and there was no significant difference in overall VAS trajectory between the subgroups (GEE: p=0.471), suggesting that there was no ceiling effect of baseline pain severity on proportionate benefit.

### Analgesic-Sparing Effect

Mean analgesic dose consumption declined from 6.38 ± 2.01 doses (median 5.00, IQR 5–7) during the Day 0–12 interval to 1.56 ± 1.96 doses (median 0.00, IQR 0–3) during the Day 36–49 interval — an overall reduction of approximately 75.5% from peak consumption (Friedman χ^2^ = 126.1, p<0.001). Table 3 shows data between intervals.

**Table 3.** Analgesic Dose Consumption Across Study Intervals (n=76).

| Interval | Mean (SD) | Median (IQR) | Range |
| --- | --- | --- | --- |
| Day 0–12 | 6.38 (2.01) | 5.00 (5–7) | 3.00–14.00 |
| Day 12–24 | 2.54 (2.91) | 2.00 (0–5) | 0.00–12.00 |
| Day 24–36 | 1.97 (2.38) | 1.00 (0–3.5) | 0.00–7.00 |
| Day 36–49 | 1.56 (1.96) | 0.00 (0–3) | 0.00–8.00 |

*Friedman test: $\chi^2 = 126.1$ , $p<0.001$ .*

### Trigger Point Burden

Mean active MTrP count remained stable from Day 0 (1.37 ± 0.59) through Day 24 (1.36 ± 0.56), with gradual reduction to 1.21 ± 0.50 at Day 36 and 1.05 ± 0.49 at Day 49. A significant reduction from baseline was first observed at Day 49 (mean absolute difference 0.32 ± 0.52; 23.4% mean reduction; p=0.031). The overall change between all timepoints was statistically significant (Friedman χ^2^ = 65.9, p<0.001).

### Safety and Tolerability

Of 76 participants, 75 (98.7%; 95% CI: 91.9–99.9%) completed the study without adverse events. One adverse event recorded was gastric irritation (Grade 2) in a male participant on Day 28 that resolved within 24 hours without drug withdrawal. On the WHO-UMC scale, causality was given a rating of Possible, NSAID intake on the previous day was noted as a likely confounding factor, and there was no recurrence on rechallenge. There were no serious adverse events reported for the entire cohort (100%; 94.0–100.0%).

## Discussion

In this exploratory pilot trial — to our knowledge the first clinical evaluation of native Himalayan Shilajit as an analgesic in a defined musculoskeletal pain condition — 73.7% of participants with MPS achieved a clinically meaningful ≥30% VAS reduction by Day 49, the IMMPACT-endorsed threshold for a minimum clinically important difference in chronic pain(36,37). The delayed onset – with no significant pain reduction at Day 12, significant reduction at Day 24, and the greatest increase in new responders at Day 49 – is a logical result of the pharmacology of Shilajit. The main active ingredients in it, fulvic acid and DBPs, are mitochondrial electron carriers that boost ATP production and restore adenylate energy charge(21,22), directly targeting the energy crisis that is at the heart of MPS pathophysiology(4,9). This bioenergetic restoration is a slow biological process by its nature. Bhattacharyya et al. demonstrated that Shilajit restored ATP levels, adenylate energy charge, and total adenine nucleotide pools in exercise-induced energy-depleted animal models, with effects comparable to CoQ10(21). The transient plateau observed between Day 24 and Day 36 may be a transitional period where peripheral anti-inflammatory activity starts to modulate central sensitization in the dorsal horn, which is eventually followed by a second wave of neurological modulation, driven by the known GABA- and glycine-mimetic activity of Shilajit at substantia gelatinosa neurons(10,11,25).

The analgesic-sparing effect is clinically important in its own right. Shilajit has been shown to have anti-ulcerogenic activity in preclinical studies similar to that of omeprazole(44), which further supports the clinical value of Shilajit as an adjunct in a population exposed to chronic analgesics. Mechanistic explanation for the analgesic-sparing effect involves the ability of fulvic acid to bind to complement, and its ability to inhibit pro-inflammatory cytokines such as tumor necrosis factor alpha (TNF-α) and interleukins(24), as well as its ability to modulate central opioid receptor function without inducing morphine tolerance, as shown by Tiwari et al(45).

Delayed trigger point resolution (significant at Day 49, well after VAS showed signs of improvements in symptoms) is indicative of the time-dependent aspects of structural musculoskeletal healing. Das et al. demonstrated that Shilajit supplementation upregulated 17 extracellular matrix (ECM) genes including collagen (up to 5.18-fold), fibronectin, fibrillin, and decorin over eight weeks, providing direct evidence for active ECM remodelling that would underpin structural resolution of trigger points(46). Supporting evidence for a delayed structural musculoskeletal effect has been provided by Keller et al. who showed a significant decrease in serum hydroxyproline, a marker of collagen degradation, with Shilajit after 8 weeks(47).

The safety profile is reassuring. With 98.7% of participants completing the study without adverse events and no serious adverse events across the cohort, and with the single Grade 2 event confounded by prior NSAID intake, our findings are consistent with chronic toxicity data from 91-day animal studies at doses up to 5000 mg/kg(48) and the absence of adverse biochemical events in human trials of up to 48-week duration(29,31). This safety advantage is particularly relevant given that NSAIDs — the standard pharmacological treatment in this population—carry well-documented risks of gastrointestinal, renal, and cardiovascular harm with chronic use(49).

There are some limitations of the study. Due to the lack of a placebo arm, it is difficult to make a definitive causal inference because of the well-documented placebo effect in chronic pain studies. There is potential for performance and detection bias for subjective outcomes in the open-label design. The sample size of 76 is appropriate for an exploratory pilot study but restricts the power of subgroups which is evident in the broad confidence intervals in the multivariable regression. Single centre design limits generalizability. The 49-day observation period does not include long-term efficacy or safety. Analgesic consumption was self-reported and recall bias is possible, and the subtherapeutic dose of the binding agent Triphala, although unlikely to be a significant factor, cannot be completely ruled out as a confounding factor. Mechanistic biomarkers which were not measured, such as inflammatory cytokines, oxidative stress indices, and mitochondrial function markers, would be useful in future trials.

In spite of these limitations, the 5.0% dropout rate is comparable with published dropout rates of 5 to 46% in chronic pain trials(50). The consistent response across diverse subgroups aligns with Shilajit’s pleiotropic pharmacology and, together with existing mechanistic and clinical evidence, supports a larger confirmatory trial. These findings justify a larger, randomized, double-blind, placebo-controlled trial incorporating biomarker endpoints and longer follow-up to definitively establish the efficacy and safety of native Himalayan Shilajit in MPS.

## Data Availability

All data produced in the present study are available upon reasonable request to the authors

## Data Availability

The datasets generated and analysed during the current study are available from the corresponding author on reasonable request.

## AUTHORS INFORMATION

### Author Contributions

DB: conceptualisation, investigation, data curation, formal analysis, and writing – original draft. PKP: conceptualisation, methodology, supervision, and writing – review and editing. RK: supervision, resources, and writing – review and editing. VSP: methodology and writing – review and editing. RY: resources, participant recruitment, and writing – review and editing. GC: pharmacovigilance oversight, methodology, and writing – review and editing. ST and DS: analytical validation of the study drug and writing – review and editing. RSK: preparation and purification of the study drug and writing – review and editing. All authors read and approved the final manuscript.

## Acknowledgements

The authors thank all participants and their families. Thanks to Swamiji Premanandji, Sivanand Ashram, Ganeshpur, Uttarkashi, India who has provided the Shilajit. We acknowledge the contribution of the Department of Biosciences and Bioengineering, IIT Roorkee, and the tablet formulation by Nagarjun Pharmacy, Dehradun. We thank the Institutional Ethics Committee and Pharmacovigilance Department of AIIMS Rishikesh for their oversight.

## References

1. Hoy D, March L, Brooks P, et al. The global burden of low back pain: estimates from the Global Burden of Disease 2010 study. Ann Rheum Dis. 2014 Jun;73(6):968–74.

2. Simons DG. Clinical and etiological update of myofascial pain from trigger points. J Musculoskelet Pain. 1996;4(1-2):93–122.

3. Jafri MS. Mechanisms of myofascial pain. Int Sch Res Notices. 2014;2014:523924.

4. Bron C, Dommerholt JD. Etiology of myofascial trigger points. Curr Pain Headache Rep. 2012 Oct;16(5):439–44.

5. Shah JP, Thaker N, Heimur J, Aredo JV, Sikdar S, Gerber L. Myofascial trigger points then and now: a historical and scientific perspective. PM R. 2015;7(7):746–61.

6. Saxena A, Chansoria M, Tomar G, Kumar A. Myofascial pain syndrome: an overview. J Pain Palliat Care Pharmacother. 2015 Mar;29(1):16–21.

7. Skootsky SA, Jaeger B, Oye RK. Prevalence of myofascial pain in general internal medicine practice. West J Med. 1989;151(2):157–60.

8. Cao QW, Peng BG, Wang L, et al. Expert consensus on the diagnosis and treatment of myofascial pain syndrome. World J Clin Cases. 2021;9(9):2077–89.

9. Shah JP, Gilliams EA. Uncovering the biochemical milieu of myofascial trigger points using in vivo microdialysis: an application of muscle pain concepts to myofascial pain syndrome. J Bodyw Mov Ther. 2008 Oct;12(4):371–84.

10. Wall PD, Woolf CJ. Muscle but not cutaneous C-afferent input produces prolonged increases in the excitability of the flexion reflex in the rat. J Physiol. 1984 Nov;356(1):443–58.

11. Fernández-de-las-Peñas C, Dommerholt J. Myofascial trigger points: peripheral or central phenomenon? Curr Rheumatol Rep. 2014 Jan;16(1):395.

12. Galasso A, Urits I, An D, et al. A comprehensive review of the treatment and management of myofascial pain syndrome. Curr Pain Headache Rep. 2020 Jun;24(8):43.

13. Rainsford KD. Profile and mechanisms of gastrointestinal and other side effects of nonsteroidal anti-inflammatory drugs (NSAIDs). Am J Med. 1999 Dec;107(6 Suppl 1):27–35.

14. Borg-Stein J, Iaccarino MA. Myofascial pain syndrome treatments. Phys Med Rehabil Clin N Am. 2014;25(2):357–74.

15. Annaswamy TM, De Luigi AJ, O’Neill BJ, Keole N, Berbrayer D. Emerging concepts in the treatment of myofascial pain: a review of medications, modalities, and needle-based interventions. PM R. 2011;3(10):940–61.

16. World Health Organization. WHO international standard terminologies on Ayurveda [Internet]. Geneva: WHO; 2022 [cited 2026 May 23]. Available from: https://www.who.int/publications/i/item/9789240064935

17. Agarwal SP, Khanna R, Karmarkar R, Anwer MK, Khar RK. Shilajit: a review. Phytother Res. 2007 May;21(5):401–5.

18. Meena H, Pandey HK, Arya MC, Ahmed Z. Shilajit: a panacea for high-altitude problems. Int J Ayurveda Res. 2010;1(1):37–40.

19. Wilson E, Rajamanickam GV, Dubey GP, et al. Review on shilajit used in traditional Indian medicine. J Ethnopharmacol. 2011 Jun;136(1):1–9.

20. Carrasco-Gallardo C, Guzmán L, Maccioni RB. Shilajit: a natural phytocomplex with potential procognitive activity. Int J Alzheimers Dis. 2012;2012:674142.

21. Bhattacharyya S, Pal D, Gupta AK, Ganguly P, Majumder UK, Ghosal S. Beneficial effect of processed shilajit on swimming exercise induced impaired energy status of mice. Pharmacologyonline. 2009;1:817–25.

22. Visser SA. Effect of humic substances on mitochondrial respiration and oxidative phosphorylation. Sci Total Environ. 1987;62:347–54.

23. Van Rensburg CEJ. The antiinflammatory properties of humic substances: a mini review. Phytother Res. 2015 Jun;29(6):791–5.

24. Schepetkin IA, Xie G, Jutila MA, Quinn MT. Complement-fixing activity of fulvic acid from shilajit and other natural sources. Phytother Res. 2009 Mar;23(3):373–84.

25. Yin H, Yang EJ, Park SJ, Han SK. Glycine- and GABA-mimetic actions of shilajit on the substantia gelatinosa neurons of the trigeminal subnucleus caudalis in mice. Korean J Physiol Pharmacol. 2011 Oct;15(5):285–9.

26. Bhattacharya SK, Sen AP, Ghosal S. Effects of shilajit on biogenic free radicals. Phytother Res. 1995 Feb;9(1):56–9.

27. Jafari M, Forootanfar H, Ameri A, et al. Antioxidant, cytotoxic and hyperalgesia-suppressing activity of a native Shilajit obtained from Bahr Aseman mountains. Pak J Pharm Sci. 2019;32(5):2167–73.

28. Stohs SJ. Safety and efficacy of shilajit (mumie, moomiyo). Phytother Res. 2014;28(4):475–9.

29. Pandit S, Biswas S, Jana U, De RK, Mukhopadhyay SC, Biswas TK. Clinical evaluation of purified Shilajit on testosterone levels in healthy volunteers. Andrologia. 2016 Jun;48(5):570–5.

30. Yadav D, Mishra S, Shah KM, Reddy ST, Gupta R, Pandey P. Safety and efficacy of TruBlk™ shilajit resin supplementation on physical performance and blood biomarkers in healthy adults: a 28-day open-label pilot study. Cureus. 2026 Jan;18(1):e102372.

31. Pingali U, Nutalapati C. Shilajit extract reduces oxidative stress, inflammation, and bone loss to dose-dependently preserve bone mineral density in postmenopausal women with osteopenia: a randomized, double-blind, placebo-controlled trial. Phytomedicine. 2022 Oct;105:154334.

32. Steen JP, Jaiswal KS, Kumbhare D. Myofascial pain syndrome: an update on clinical characteristics, etiopathogenesis, diagnosis, and treatment. Muscle Nerve. 2025 May;71(5):889–910.

33. Simons DG, Travell JG, Simons LS. Myofascial pain and dysfunction: the trigger point manual. Vol. 1. 2nd ed. Baltimore: Williams & Wilkins; 1999. 1056 p.

34. Randhawa GK. Cow urine distillate as bioenhancer. J Ayurveda Integr Med. 2010 Oct;1(4):240–1.

35. Singh S, Tripathi JS, Rai NP. An appraisal of the bioavailability enhancers in Ayurveda in the light of recent pharmacological advances. Ayu. 2016;37(1):3.

36. Farrar JT, Young JP, LaMoreaux L, Werth JL, Poole RM. Clinical importance of changes in chronic pain intensity measured on an 11-point numerical pain rating scale. Pain. 2001;94(2):149–58.

37. Dworkin RH, Turk DC, Wyrwich KW, et al. Interpreting the clinical importance of treatment outcomes in chronic pain clinical trials: IMMPACT recommendations. J Pain. 2008 Feb;9(2):105–21.

38. Haefeli M, Elfering A. Pain assessment. Eur Spine J. 2006 Jan;15(Suppl 1):S17–24.

39. Carlsson AM. Assessment of chronic pain. I. Aspects of the reliability and validity of the visual analogue scale. Pain. 1983;16(1):87–101.

40. Julious SA. Sample size of 12 per group rule of thumb for a pilot study. Pharm Stat. 2005 Oct;4(4):287–91.

41. Lancaster GA, Dodd S, Williamson PR. Design and analysis of pilot studies: recommendations for good practice. J Eval Clin Pract. 2004 May;10(2):307–12.

42. Thabane L, Ma J, Chu R, et al. A tutorial on pilot studies: the what, why and how. BMC Med Res Methodol. 2010;10:1.

43. Boonstra AM, Preuper HRS, Balk GA, Stewart RE. Cut-off points for mild, moderate, and severe pain on the visual analogue scale for pain in patients with chronic musculoskeletal pain. Pain. 2014;155(12):2545–50.

44. Goel RK, Banerjee RS, Acharya SB. Antiulcerogenic and antiinflammatory studies with shilajit. J Ethnopharmacol. 1990 Apr;29(1):95–103.

45. Tiwari P, Ramarao P, Ghosal S. Effects of shilajit on the development of tolerance to morphine in mice. Phytother Res. 2001;15(2):177–9.

46. Das A, Datta S, Rhea B, et al. The human skeletal muscle transcriptome in response to oral shilajit supplementation. J Med Food. 2016 Jul;19(7):701–9.

47. Keller JL, Housh TJ, Hill EC, Smith CM, Schmidt RJ, Johnson GO. The effects of shilajit supplementation on fatigue-induced decreases in muscular strength and serum hydroxyproline levels. J Int Soc Sports Nutr. 2019 Feb;16(1):3.

48. Velmurugan C, Vivek B, Wilson E, Bharathi T, Sundaram T. Evaluation of safety profile of black shilajit after 91 days repeated administration in rats. Asian Pac J Trop Biomed. 2012;2(3):210–4.

49. Baigent C, Bhala N, Emberson J, et al. Vascular and upper gastrointestinal effects of non-steroidal anti-inflammatory drugs: meta-analyses of individual participant data from randomised trials. Lancet. 2013;382(9894):769–79.

50. Oosterhaven J, Wittink H, Mollema J, Kruitwagen C, Devillé W. Predictors of dropout in interdisciplinary chronic pain management programmes: a systematic review. J Rehabil Med. 2019;51(1):2–10.

